# Rapid And high throughput RT-qPCR assay for identification and differentiation between SARS-CoV-2 variants B.1.1.7 and B.1.351

**DOI:** 10.1101/2021.05.19.21257439

**Authors:** Oran Erster, Ella Mendelson, Virginia Levy, Areej Kabat, Batya Mannasse, Hadar Asraf, Roberto Azar, Yaniv Ali, Rachel Shirazi, Efrat Bucris, Dana Bar-Ilan, Orna Mor, Michal Mandelboim, Danit Sofer, Shai Fleishon, Neta S Zukerman

## Abstract

Emerging SARS-CoV-2 (SC-2) variants with increased infectivity and vaccine resistance are of major concern. Rapid identification of such variants is important for the public health activities and provide valuable data for epidemiological and policy decision making. We developed a multiplex quantitative RT-qPCR (qPCR) assay that can specifically identify and differentiate between the emerging B.1.1.7 and B.1.351 SC-2 variants. In a single assay, we combined four reactions: one that detects SC-2 RNA independently of the strain, one that detects the D3L mutation, which is specific to variant B.1.1.7, and one that detects the 242-244 deletion, which is specific to variant B.1.351. The fourth reaction identifies human RNAseP gene, serving as an endogenous control for RNA extraction integrity. We show that the strain-specific reactions target mutations that are strongly associated with the target variants, and not with other major known variants. The assay’s specificity was tested against a panel of respiratory pathogens (n=16), showing high specificity towards SC-2 RNA. The assay’s sensitivity was assessed using both *In-vitro* transcribed RNA and clinical samples, and was determined to be between 20 and 40 viral RNA copies per reaction. The assay performance was corroborated with Sanger and whole genome sequencing, showing complete agreement with the sequencing results. The new assay is currently implemented in the routine diagnostic work at the Central Virology Laboratory, and may be used in other laboratories to facilitate the diagnosis of these major worldwide circulating SC-2 variants.

## INTRODUCTION

The recent emergence of new SARS-COV-2 (SC-2) variants of concern (VOC) B.1.1.7 in the UK and B.1.351 in South Africa, both characterized by increased transmissibility and potential vaccine resistance (**1-4**), prompted dedicated surveillance by Israel’s Central Virology Laboratory, to monitor their incursion into Israel.

Since the genome of variant B.1.1.7 contains 23 unique mutations, and that of variant B.1.351 contains 18 unique mutations, compared with the original Wuhan strain, it is practically impossible to detect all of them in one qPCR assay. Thus far, the Thermo Fisher SC-2 detection kit (CAT CCU002, https://www.thermofisher.com) was utilized to identify B.1.1.7 suspected samples (**3)**. One of the reactions in this kit is directed to the viral Spike gene, and is negative when the template sequence contains the 69-70 deletion, which is one of the B.1.1.7 variant mutations. However, this deletion was also detected independently, is not unique to the B.1.1.7 variant, and can therefore often be misleading. Moreover, the absence of the S reaction in this assay may result from inhibition of this reaction, and therefore may not necessarily indicate the presence of the deletion. Variant B.1.1.7 was first reported in the UK on September 2020 and by December 2020 became the dominant strain in the country (**2**). Its increased infectivity led to its rapid spread, with severe consequences on public health and global economy (**4**). Likewise, from its first detection in Israel on December 23, this variant now comprises over 90% of the positive cases (O. Erster and N. Zuckerman, Unpublished data).

An additional SC-2 VOC is the B.1.351 variant, which contains both B.1.351-unique mutations and mutations that are present in other notable VOC, such as the SGF deletion in the nsp6 gene, and the N501Y substitution in the spike gene that is identified also in the B1.1.7 **(5**). Like variant B.1.1.7, this variant was found to be more infectious than the WT strain. *In vitro* studies also show that it has increased resistance to serum of recovered and vaccinated patients, thereby posing a serious threat on the efficacy of current vaccination campaigns (**6-8**).

The emergence of these two variants, as well as other recently emerging SC-2 strains, necessitates constant improvement of the diagnostic tools used to combat the COVID-19 pandemic. In addition to performing a rapid, sensitive and specific detection of the viral RNA, diagnostic tests are now required to differentiate between circulating strains to provide valuable epidemiological data for policy decision making. Commercial assays developed recently (, http://www.kogene.co.kr/eng/sub/product/covid-19/) detect specific key mutations. However, recent studies showed that in spite of sharing such mutations, variants might differ in their antibody resistance capacity, prompting development of variant specific assays [**5, 9**]. These findings highlight the need to determine the identity of circulating strains, not only specific mutations. By designing a multiplex PCR assay that positively detects the presence of unique mutations that are strongly associated with variants B.1.1.7 and B.1.351, rapid, economical high throughput screening can be performed, enabling robust and specific identification of these variants in SC-2 positive clinical samples.

In this report, we describe a differential RT-qPCR assay that detects the presence of mutations strongly associated with variants B.1.1.7 and B.1.351. We demonstrate that implementation of this novel multiplex assay allows a sensitive and specific detection of SC-2 RNA, with the advantage of variant differentiation in a single assay.

## MATERIALS AND METHODS

### Design of VOC-specific qPCR reactions

Analysis of SC-2 sequences and primer simulations were performed using the Geneious software package (https://www.geneious.com/) and the NCBI BLAST analysis tools (https://blast.ncbi.nlm.nih.gov/). SC-2 sequences were obtained from GISAID initiative website (https://www.gisaid.org/) and Analyzed using the Geneious software. All primer and probe sequences are detailed in **Table 1**.

**Table 1.**
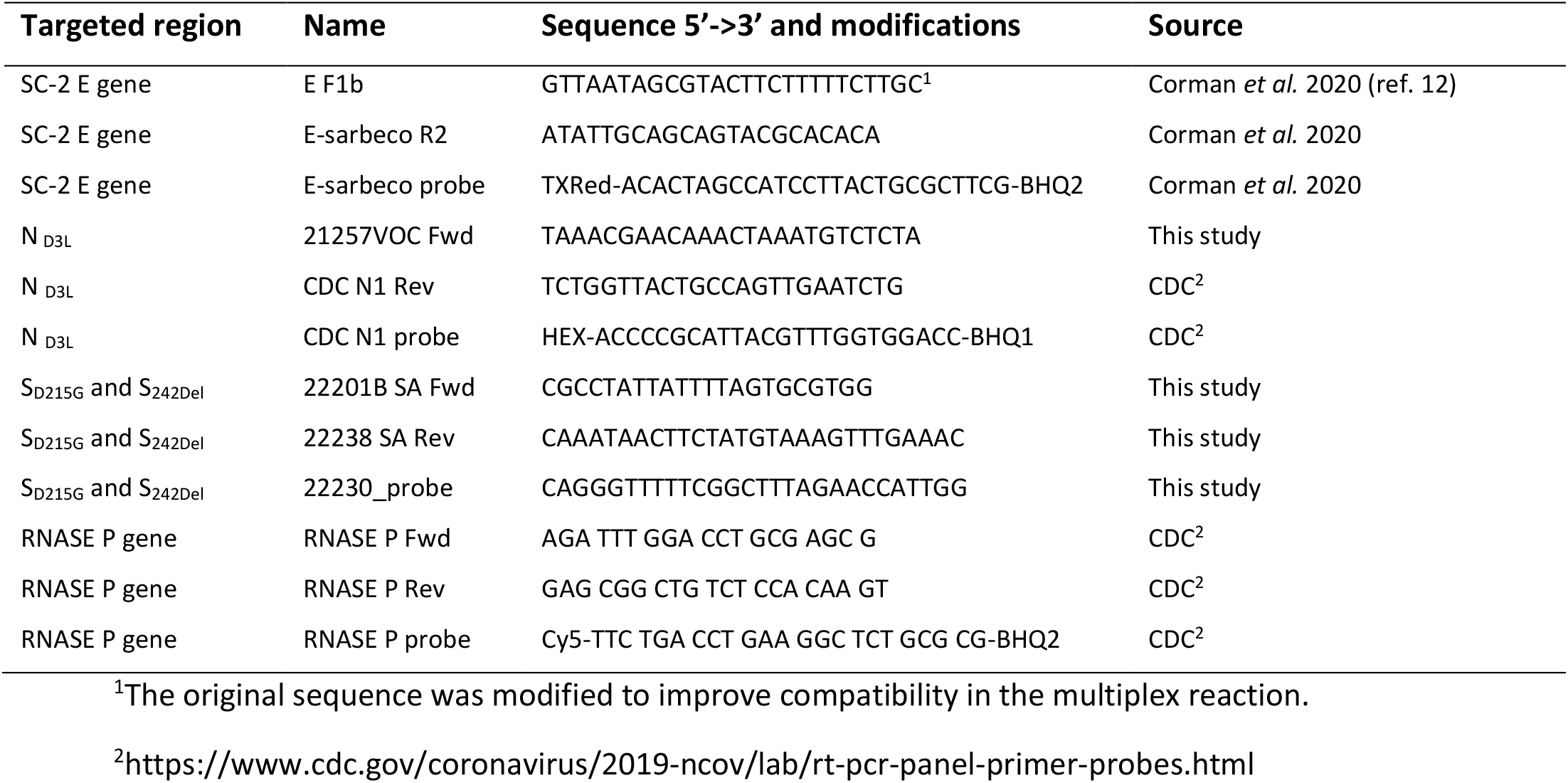
Primers and probes for the differential detection reaction design.

### Processing of SC-2 clinical samples

Nasopharyngeal swab samples suspected to contain SC-2 in viral transport media (VTM) were inactivated by heating at 70°C for 30 minutes, or, if intended to be used for culturing, inactivated by addition of 200µl lysis buffer (Zymo research, https://www.zymoresearch.com/products/dna-rna-lysis-buffer) to 200µl VTM.. Total RNA extraction was performed with either the Roche MagNA pure 96 system (https://lifescience.roche.com/), or the PSS MagLEAD instrument (http://www.pss.co.jp/english/). The eluted RNA was stored at -80°C for further use, or used immediately thereafter for the PCR test.

### Design and synthesis of In vitro transcribed standard RNA segments

In order to establish the analytical Limit of Detection (LOD) and obtain standard controls for WT and mutant target sequences, genomic regions including the E, S_242_, S_RBD_, N and RNASE P were amplified using primers that contain the T7 promoter minimal sequence (**Table 2**) with the MyTaq One-step RT-PCR kit. The resulting PCR products were transcribed *In vitro* to RNA using the T7 Megascript kit according to the manufacturer’s instructions (Thermo Fisher, https://www.thermofisher.com/). The *In-vitro* transcribed RNA was purified and its concentration was determined using Nanodrop spectrophotometer and stored at -80°C.

**Table 2.**
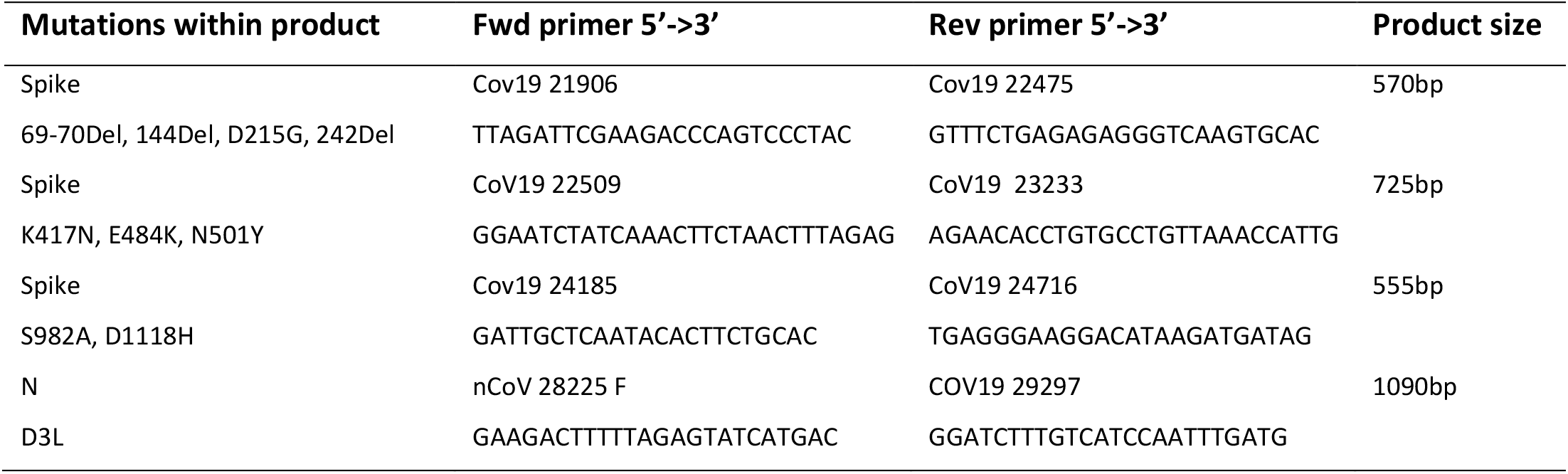
Cloning reactions design.

### RT-qPCR

RT-qPCR mix was prepared with the Meridian (formerly Bioline) SensiFast one-step mix (https://www.bioline.com/). Initial optimization was performed by setting an annealing temperature gradient. Following optimization of the reaction conditions, the final mix concentration was determined. The reaction mix was assembled as follows: SensiFast Probe Lo-ROX One-Step: 12.5µl, E Sarbeco-F1b: 400nM, E-Sarbeco-R: 400nM, E-Sarbeco Probe: 200nM, 21257VOC Fwd: 750nM, CDC N1 Rev: 750nM, CDC N1 probe: 300nM, 22201B SA Fwd: 600nM, 22238 SA Rev: 800nM, 22230 probe: 300nM, RNASE P Fwd: 300nM, RNASE P Rev: 300nM, RNASE P probe: 200nM, RT Enzyme: 0.2µl, RNAse inhibitor: 0.2µl, ddH_2_O – to a final volume of 1µl.

The amplification was performed in Bio-Rad CFX96 thermal cycler (https://www.bio-rad.com) using the following conditions:

(a) 45°C for 10’, (b) 95°C for 2’:20’’, 45X[(C) 95°C for 4’’, (d) 62.2°C for 28’’]. Fluorescence was recorder at each cycle during the annealing and extension step (62.2°C for 28’’). The reaction data were analyzed using the Bio-Rad CFX Maestro software.

### Sanger Sequencing of suspected samples

In order to identify mutations of the variants of interest in suspected samples, several rapid sequencing reactions of the Spike gene were designed and implemented. This enabled the first identification of the B.1.1.7-related mutations 69-70Del, 144Del, N501Y, S982A and D1118H, and B.1.351-related mutations D215G, 242Del K417N, N501Y and E484K. Primer sets for the rapid sequencing reactions of the Spike and Nucleocapside (N) genes are detailed in **Table 2**.

Endpoint PCR reactions were performed with MyTaq One-step RT-PCR kit (Meridian, https://www.bioline.com/mytaq-mix.html) according to the manufacturer’s instructions. Resulting PCR products were analyzed using agarose gel electrophoresis and sequenced using the ABI 3500 Bioanalyzer.

### Next Generation Whole genome sequencing of clinical samples

COVID-seq kit was used for library preparation as per manufacturer’s instructions (Illumina, https://www.illumina.com/). Library validation and mean fragment size was determined by TapeStation 4200 via DNA HS D1000 kit (Agilent, https://www.agilent.com/). Libraries were pooled, denatured and diluted to 10pM and sequenced on NovaSeq (Illumina).

### Bioinformatic and phylogenetic analysis

Fastq files underwent quality control using FastQC (www.bioinformatics.babraham.ac.uk/projects/fastqc/) and MultiQC [**10**] and low-quality sequences were filtered using trimmomatic [PMID: 24695404]. Mapping to SARS-CoV-2 genome (NC_045512.2) was performed with BWA mem [PMID: 19451168]. SAMtools suite [PMID: 19505943] was used to filter unmapped reads, sort and index bam files. Consensus fasta sequences were constructed for each sample using iVar (https://andersen-lab.github.io/ivar/html/manualpage.html), with Ns inserted in positions with sequencing depth lower than five. Sequences were aligned with the SARS-CoV-2 reference sequence (NC_045512.2) with MAFFT [**11**] and mutation analysis was done with a custom R code using Bioconductor package seqinr [citation info in: https://cran.r-project.org/web/packages/seqinr/citation.html].

## RESULTS

### Development of differential COV19 VOC RT-qPCR

Analysis of mutations characteristic of variants B.1.1.7 and B.1.351 showed that some of them were mutual to both, like the SGF deletion in nsp6 (positions 11285-11294 in sequence NC_045512), or the N501Y substitution in the spike protein gene (nucleotide position 23,094 in sequence NC_045512). Others, such as the 69-70 deletion and N501Y substitution, developed independently and were detected in samples not classified as variant B.1.1.7 (**Supplementary Figure S1**). On the other hand, the N gene D3L mutation is strongly associated with variant B.1.1.7 and is not associated with other currently dominant variants. Likewise, the D215G mutation and the S gene 242-244 deletion are strongly associated with variant B.1.351 and were not identified in other variants thus far (**Supplementary Figure S2**.). Therefore, these two regions were selected for the multiplex reactions design.

### Design of variant B.1.1.7 specific reaction

The 5’p region of variant B.1.1.7 SC-2 N gene contains a complete codon substitution, translated into D3L a.a. substitution. Additionally, there is a single “A” deletion in this region, 5 bases upstream to the N gene start codon (**Figure 1**). A selective primer was designed accordingly, to specifically detect the mutated variant. The reverse primer and the probe were based on the CDC N1 reaction (https://www.cdc.gov/coronavirus/2019-ncov/lab/rt-pcr-panel-primer-probes.html), as detailed in **Table 1**. The B.1.1.7-specific reaction was combined with an inclusive reaction based on the E-sarbeco qPCR described by Corman *et al*. (**12**), with minor modifications, as detailed in **Table 1**. This reaction was designed to detect all known CoV19 clades, thereby serving as an indicator for the presence of SC-2 RNA in the examined sample. The combined assay was then tested using WT and sequence-verified samples identified as belonging to lineage B.1.1.7 (**Figure 1**). All samples identified as belonging to clade B.1.1.7 were detected using the new multiplex assay. WT samples were either negative for the N reaction, or gave a very weak signal, approximately 10-15 cycles apart from the E reaction signal (**Figure 1**). This reaction was termed N gene D3L reaction.

**Figure 1.**
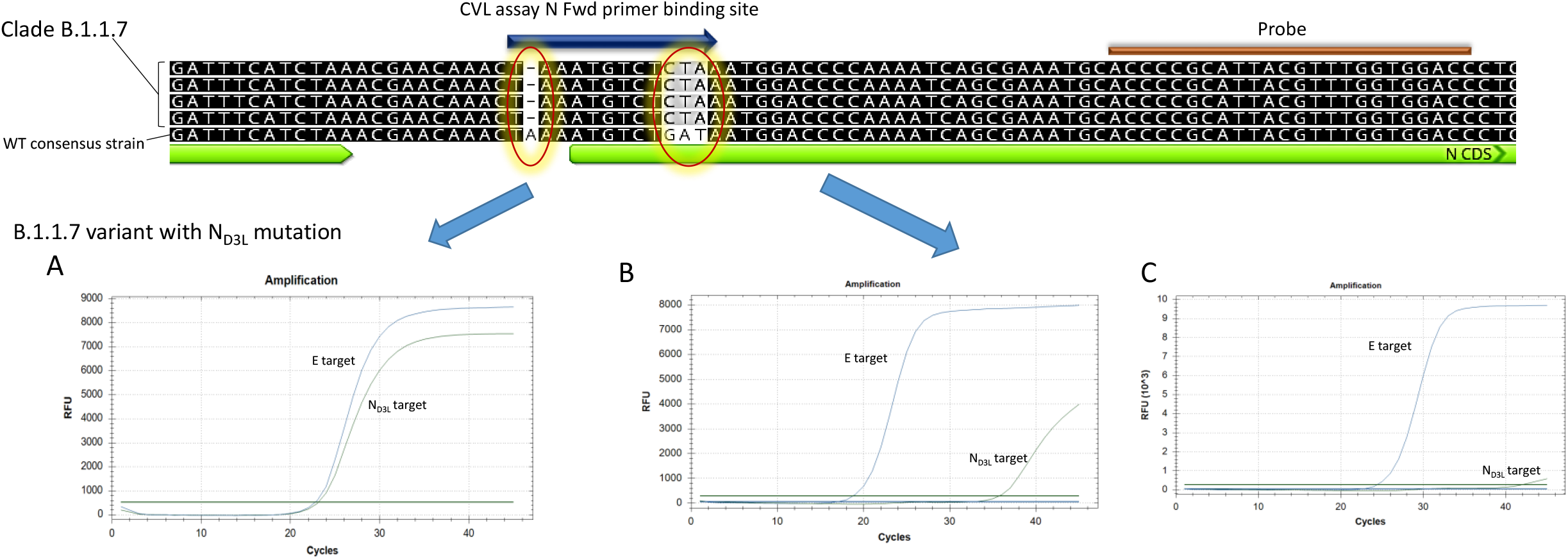
Design of variant B.1.1.7-specific reaction. A specific primer was designed based on the single nucleotide insertion and the codon substitution in the 5’p of the SC-2 N gene, both circled and highlighted. The specific “UK VOC” reaction was combined with an inclusive E-sarbeco reaction (Corman et al. 2020) that detects all SC-2 variants. The presence of the B.1.1.7 RNA in the assay results in two clear amplification curves (A). A sample containing a non-B.1.1.7 variant sequence results in a faint amplification, with a Cq different larger than eight between the E and the N curves (B), or a negative signal for the N reaction (C).

### Design of variant B.1.351 specific reaction

This variant contains two unique mutations in the spike gene, D215G and a deletion at amino acid position 242 (nucleotide positions 22281 and 22289 in sequence NC_045512). A specific reaction was developed, based on these mutations, which detects samples of lineage B.1.351, as described. The forward primer was designed to anneal to the substituted nucleotide at its 3’p end. In order to increase the primer selectivity, a mismatched base was inserted 10 b.p. downstream to the 5’p end. The reverse primer was designed to complement the region of the 242-244 deletion (**Figure 2**). This reaction resulted in a negative, or a very weak signal, when testing WT samples, and a clear signal when testing samples previously sequenced and identified as belonging to the B.1.351 clade (**Figure 2**). This reaction was then termed S_242_ reaction.

**Figure 2.**
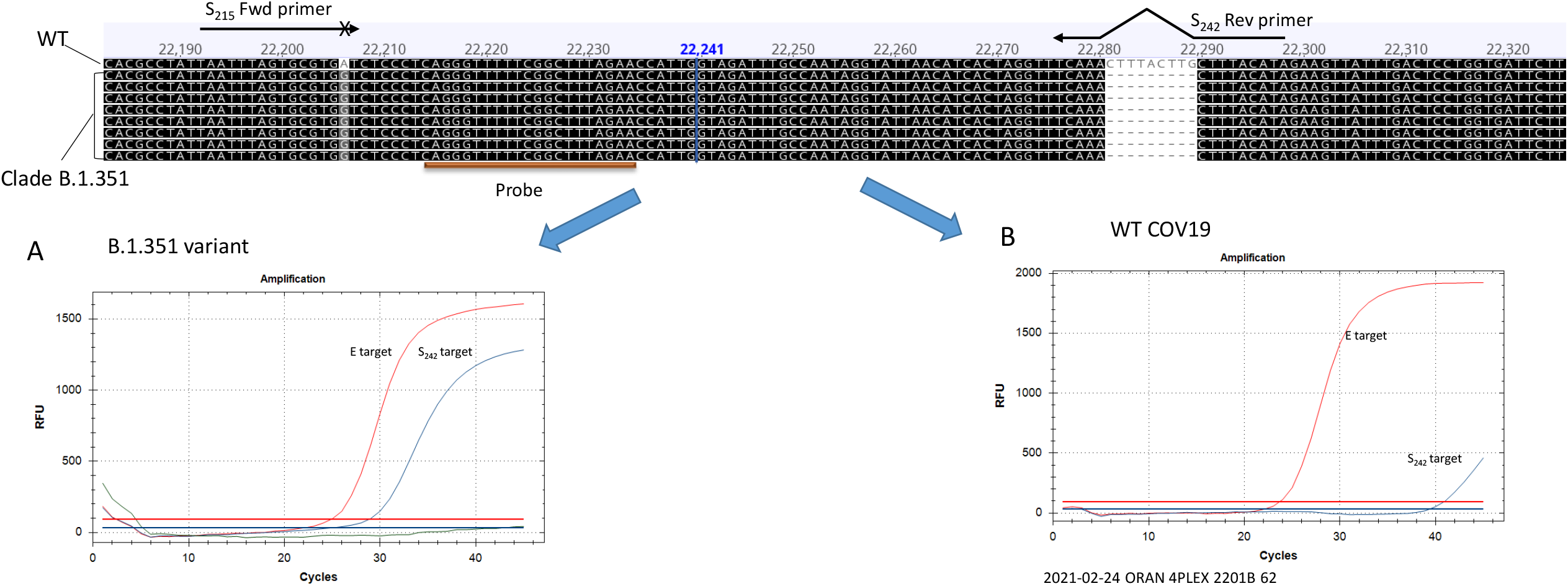
Design of variant B.1.351-specific reaction. Two primers were designed, to specifically complement the sequences containing the D215G mutation, and the 242-244 deletion. The resulting reaction gives a clear signal of the S_242_ reaction with sample belonging to clade B.1.351 (A), and a negative, or a faint signal with a WT sample (B). The inclusive E-sarbeco reaction detects the presence of both the WT and clade B.1.351 samples.

### Generation and evaluation of a multiplex SC-2 VOC reaction

The two reactions were combined with the E target reaction and a reaction targeting the RNAse P gene (https://www.cdc.gov/coronavirus/2019-ncov/lab/rt-pcr-panel-primer-probes.html) that serves as an endogenous control for RNA extraction. The multiplex reaction was then tested for sensitivity using serial dilutions of *in vitro-*transcribed RNA containing the target sequences of all four reactions. As detailed in **Supplementary Figure S3**, the calculated analytical limit of detection for the three viral targets was as follows: 29 copies per reaction for the E-sarbeco reaction, 26 copies per reaction for the N_D3L_, and 39 copies per reaction for the S_242_ reactions. This sensitivity was accomplished using a reaction that takes less than 70 minutes, thereby shortening the entire time from sample to answer. Specificity test performed with a panel of respiratory pathogens showed no cross-detection of pathogens other than SC-2, except for SARS CoV-1 (SC-1), which was detected, as expected, by the E-sarbeco reaction. Nucleic acid samples of the following pathogens were tested: *Chlamydia pneumonia*, Rhinovirus, Influenza A, Influenza B, Swine influenza, RSV A, RSV B, Parainfluenza 3, HMPV, Adenovirus, CoV OC43, CoV 229E, CoV HKU1, CoV NL63, SARS-1 and MERS CoV.

The multiplex SC-2 VOC assay was then tested using sequence-verified samples, and successfully detected all samples in full agreement with the sequencing results, thereby confirming the accuracy of the assay. Notably, in samples with high viral RNA concentration, a weak signal of S_242_ variant was observed. However, the Cq difference between the E gene target signal and the S_242_ reaction clearly indicated that the signal does not reflect the presence of the D215G and 242-244 deletion mutations, but rather a low-affinity binding of the primers to the RNA.

### Confirmatory Sequencing of suspected samples

The first three samples suspected as variant B.1.1.7 (numbered 5824, 6021 and 7075) were negative for the S reaction in the Thermo-Fisher SARS-COV-2 test, were subjected to further examination in the Israel Central Virology Laboratory (CVL). Focused Sanger sequencing of the S gene region of samples 5824 and 6021 showed the presence of the 69-70 deletion, but the B.1.1.7 variant-associated mutations 144del and N501Y substitution were absent. They both contained, however, a unique substitution - N439K, which is not associated with the B.1.1.7 variant (**Supplementary figure S4**). Sanger sequencing of the spike gene region from sample 7075, contained the following mutations, all of which characteristic of the B.1.1.7 variant: 69-70 deletion, 144 deletion, N501Y, S982A and D1118H (**Supplementary figure S5**). Subsequent complete genome analysis via next generation sequencing (NGS) of that sample confirmed the presence of all the defining mutations of the B.1.1.7 variant, thereby corroborating the initial results of the Sanger sequencing. This was the first known case of the B.1.1.7 variant in Israel.

Samples suspected as the B.1.351 variant were initially examined by Sanger sequencing of two regions of the spike protein: the 242-244 deletion, which is unique to this variant, and the receptor binding domain (RBD, nucleotide positions 22,493-23,218). Out of 20 suspected samples, four contained the following mutations in the spike gene: D215G, 242-244 deletion, K417N, E484K, and N501Y, all associated with the B.1.351 variant (**Supplementary figure S6**). Subsequent complete genome analysis via NGS confirmed the presence of the B.1.351 defining mutations.

To confirm the accuracy of the multiplex reaction, complete genome sequencing of 122 clinical samples was performed, with complete agreement with the qPCR results. For variant B.1.1.7, over 1,000 samples were examined by both the new qPCR assay and by whole genome sequencing, with complete match. These results demonstrate that the new multiplex assay described herein can be used as a rapid and reliable approach for primary classification of SC-2 B.1.1.7 and B.1.351 variants.

## DISCUSSION

The emergence of new, more contagious and potentially antigenically different SC-2 lineages pose an urgent need to adjust rapid detection methods to meet public health related needs. To meet these needs, we developed a multiplex RT-qPCR assay that can distinguish between three SC-2 lineages. The assay is rapid (∼1 hour PCR assay time) and is suitable for high throughput rapid screening. This is in contrast to Sanger sequencing or NGS, which are more informative, but are far more expensive, take significantly more time, and cannot be scaled up easily. Since the beginning of the COVID-19 pandemic, several rapid tests detecting the presence of SC-2 RNA or proteins were implemented in wide scale testing (**13**). However, the capacity to distinguish between different lineages in a time scale of hours is currently possible only using qPCR.

SC-2 variant B.1.1.7 contains numerous synonymous and non-synonymous mutations, of which the Spike gene mutations 69-70del, N501Y, and P681H received most attention due to their potential effect on virus infectivity (**14-15**). For diagnostic purposes, however, the N501Y mutation is not variant-specific, as it was identified in several variants other than variant B.1.1.7, such as B.1.351 and the P.1 variant first discovered in Brazil (**16**). The D3L substitution in the N gene used in our assay is specific to variant B.1.1.7 and was not reported in other major SC-2 lineages. Although this mutation can occur independently from other characteristic mutations, such as N501Y, its presence strongly suggests that the examined sample is the B.1.1.7 variant. Likewise, the reaction that identifies the variant B.1.351 targets mutations that are strongly associated with this variant: D215G and the triple deletion of amino acids 242-244 (**Supplementary Figure S4**). The combinations of these two reactions therefore provides a reliable tool to identify each of these two variants with high confidence. In order to increase the range of the new assay, the SC-2 inclusive reaction targeting the E gene **(12**) was combined, thereby enabling detection of the viral RNA independently of the strain examined.

A few commercial kits partially address the detection of these SC-2 variants, but they target general mutations, such as N501Y and E484K in the spike protein, and not variant-specific mutations, such as the N protein D3L substitution or the spike protein 242-244 deletion (https://www.seegene.com/assays/rp, http://www.kogene.co.kr/eng/sub/product/covid-19.asp).

The emergence of novel SC-2 variants with increased infectivity and increased resistance to current vaccines may significantly impair global large-scale detection and vaccination efforts **(17**). Moreover, it has been shown that different variants having some identical mutation in the spike coding sequence still have different infectivity and vaccine resistance characteristics due to their different set of additional mutations [**5**,**9**, Mandelboim M., unpublished data]. As a result, the pressing need to improve detection methods accordingly requires constant adjustments. Such a diagnostic tests should not only detect the presence of the viral RNA with high specificity and sensitivity, but also provide information on variant identity. Additional consideration is the relatively high cost of commercial kits, and the need to perform complex interpretation of the results, to determine the possible sample identity. The execution and analysis of the assay described herein are simple, and relatively inexpensive, compared with current commercial kits. Implementation of molecular assays such as our multiplex qPCR assay will improve SC-2 diagnosis and contribute to the ongoing efforts to contain the COVID-19 pandemic.

## Data Availability

All raw qPCR results and whole genome sequencing data are stored in the central virology laboratory and available upon request.

## ACKNOWLEDGEMENTS

The authors wish to thank the members of the Israel Central Virology Laboratory for their valuable help in this work.

## CONFLICT OF INTERESTS

The authors declare no conflict of interests.

## Notes

### Competing Interest Statement

The authors have declared no competing interest.

### Funding Statement

This study was funded by the CVL internal resources.

### Author Declarations

All CoV19-related studies in the CVL are conducted under ethical approval no. 7045-20-SMC from the Sheba Medical Center ethics committee. since all samples used in this study were anonymized leftover from clinical samples, no further approvals were required.

