## Supplemental Material File for "Rapid And high throughput RT-qPCR assay for identification and differentiation between SARS-CoV-2 variants B.1.1.7 and B.1.351"

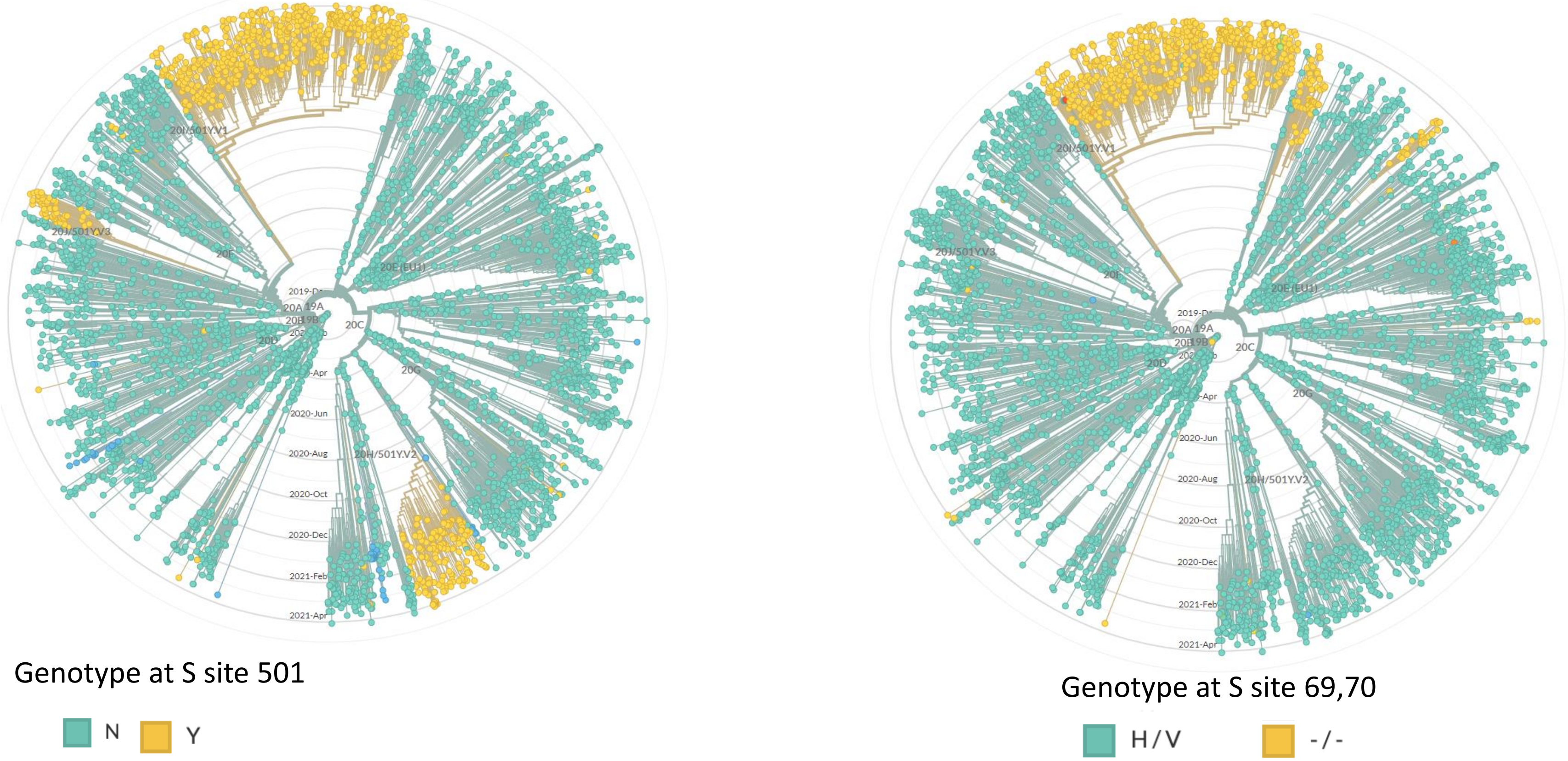


**<u>Supplemental Figure S1</u>. Global phylogenetic alignment of SARS-COV-2 lineages, showing the Prevalence of N501Y (left) and 69-70del (right) mutations.** The WT sequence (N for the 501 plot and H/V for the 69-70 plot) is colored in grey. The mutation (Y for the 501 plot and -/- for the 69-70 plot) is highlighted in yellow. Lineage annotations: 20I/501Y.V1 – B.1.1.7, 501.V2, 20H/501Y.V2 -B.1.351, 20J/501Y.V3 – P1 . Plots were generated using the Nextstrain website (https://nextstrain.org/ncov/).

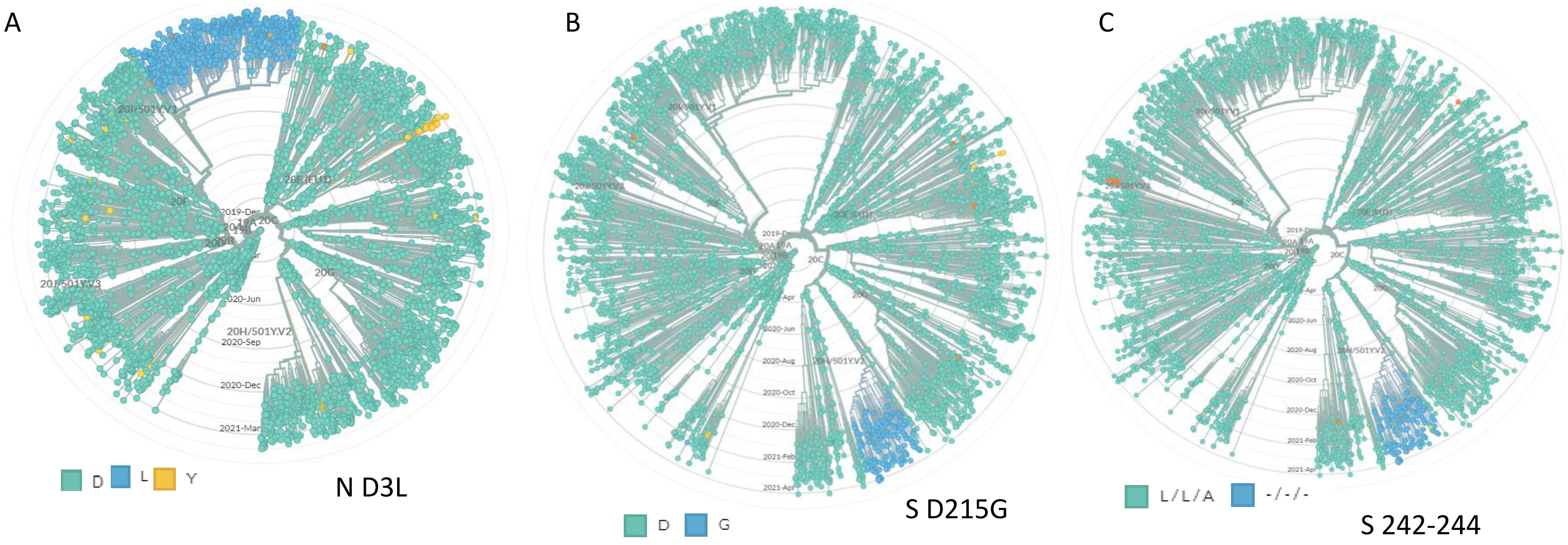


<u>**Supplemental Figure S2**</u>. **Global phylogenetic alignment of SARS-COV-2 lineages, showing the Prevalence of N (nucleocapsid), Spike (S) D215G and S 242-244del mutations.** (A) Global alignment of SC-2 showing the different mutations in position 3 of the N gene. The L substitution is colored in purple. (B) Global alignment of showing the presence of the S D215G mutation, highlighted in purple. (C) Global alignment of showing the presence of the S 242-244 deletion. The WT sequence (D for the N D3L plot, D for the D215G plot and L/L/A for the 242-244 plot) is colored in grey. Lineage annotations: 20I/501Y.V1 – B.1.1.7, 501.V2, 20H/501Y.V2 -B.1.351, 20J/501Y.V3 – P1. Plots were generated using the Nextstrain website (https://nextstrain.org/ncov/).

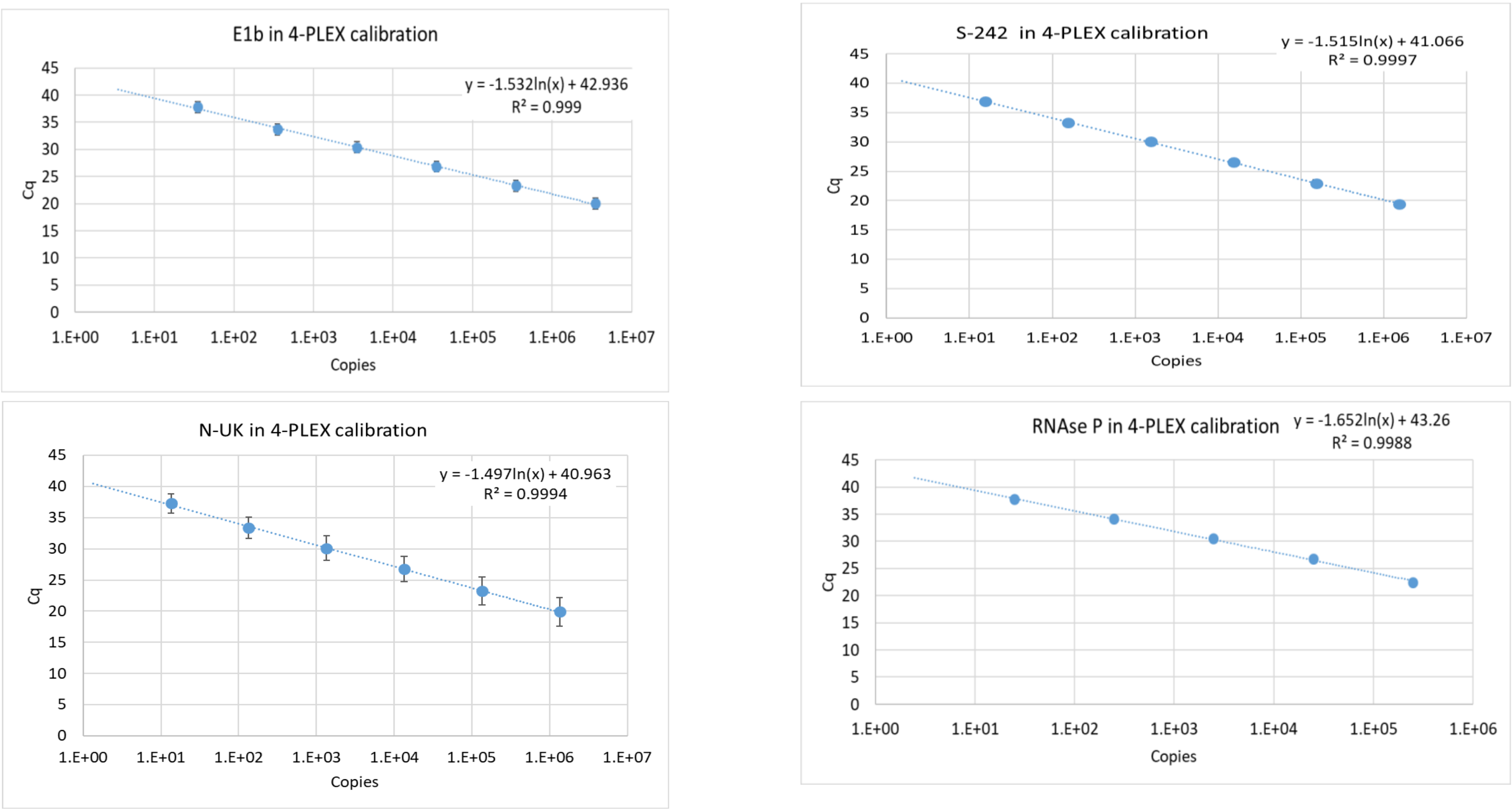


**Supplemental Figure S3. Calibration curves of the multiplex assay reactions.** *In-vitro* transcribed RNA corresponding to the target sequences was serially diluted and tested in triplicates. The average Cq values for each concentration were plotted against the calculated RNA copies. The standard deviation are of three separate repeats for each concentration. The resulting calibration formula and the $R^2$ value of the regression line are detailed in the inset of each graph.

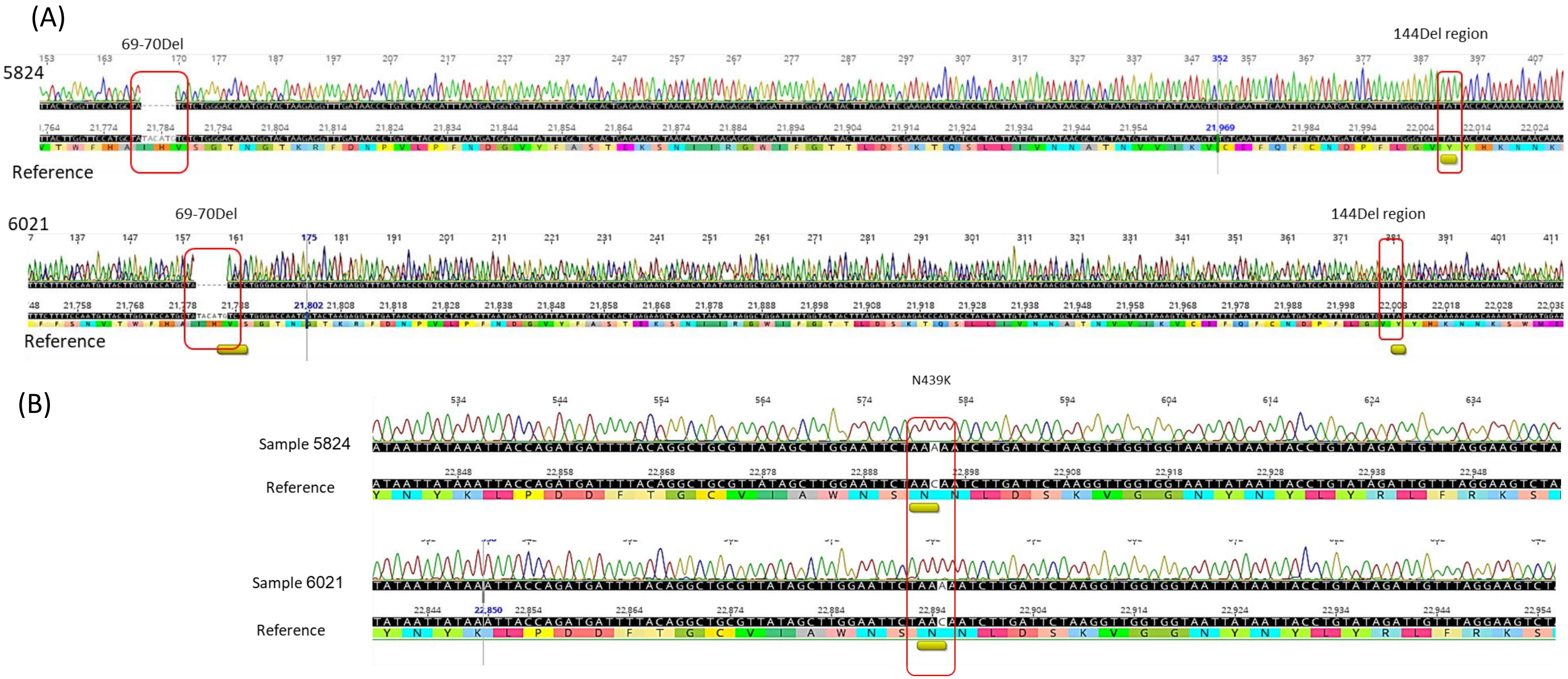


**<u>Supplemental Figure S4</u>. Alignment of lineage B.1.1.7 suspected samples with reference sequence NC_045512**. In samples 5824 and 6021, The deletion of 69-70 was evident, but the 144 deletion was absent (A). Both sample did not contain any characteristic mutations of lineage B.1.1.7 in the RBD, but did contained the N439K mutation, which is not a defining mutation of variant B.1.1.7 (B). Mutations are marked with a rectangle and an underline under the sequence. Reference sequence: accession NC_045512.

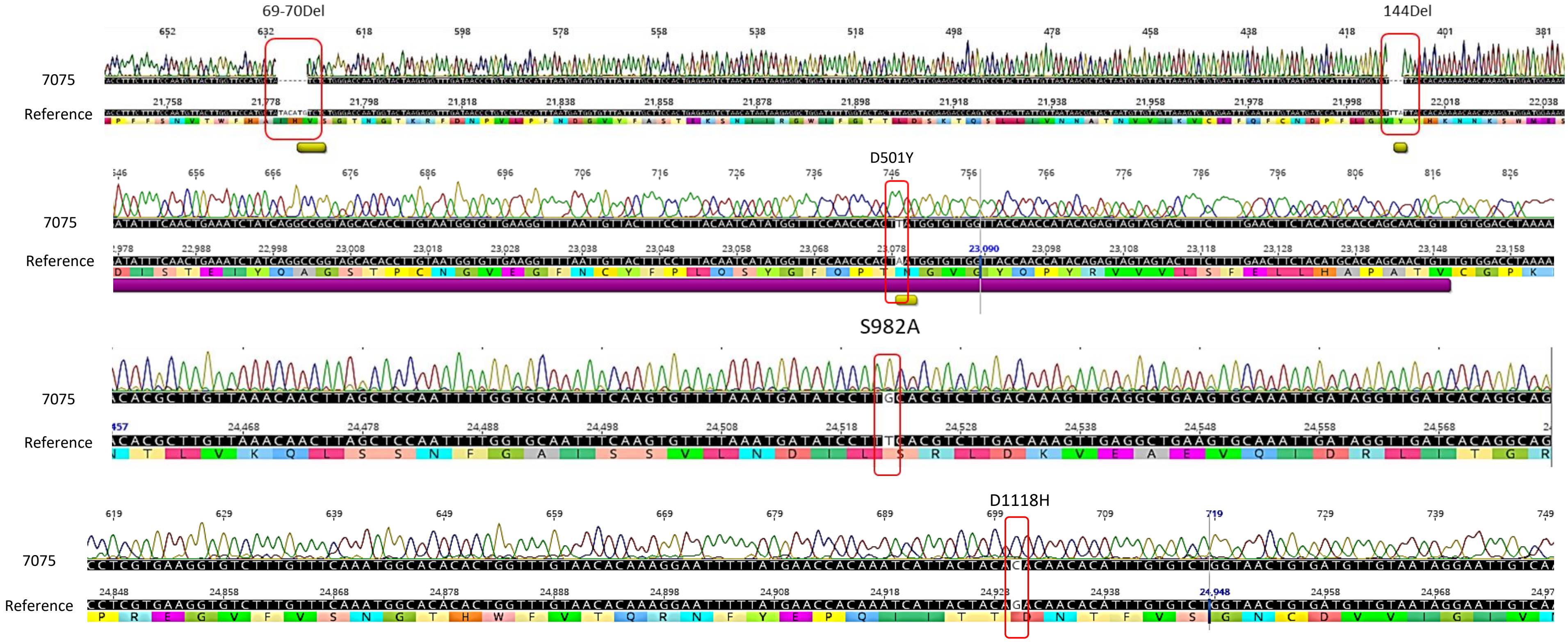


**<u>Supplemental Figure S5</u>. Sequencing of the Spike gene N-terminal and RBD regions of sample 7075**. The following Variant B.1.1.7- associated mutations were detected: 69-70 deletion, 144 deletion, D501Y, S982A and D1118H. Mutations are marked with a rectangle and an underline under the sequence. Reference sequence: accession NC_045512. Violet underline in the second alignment marks the RBD region.

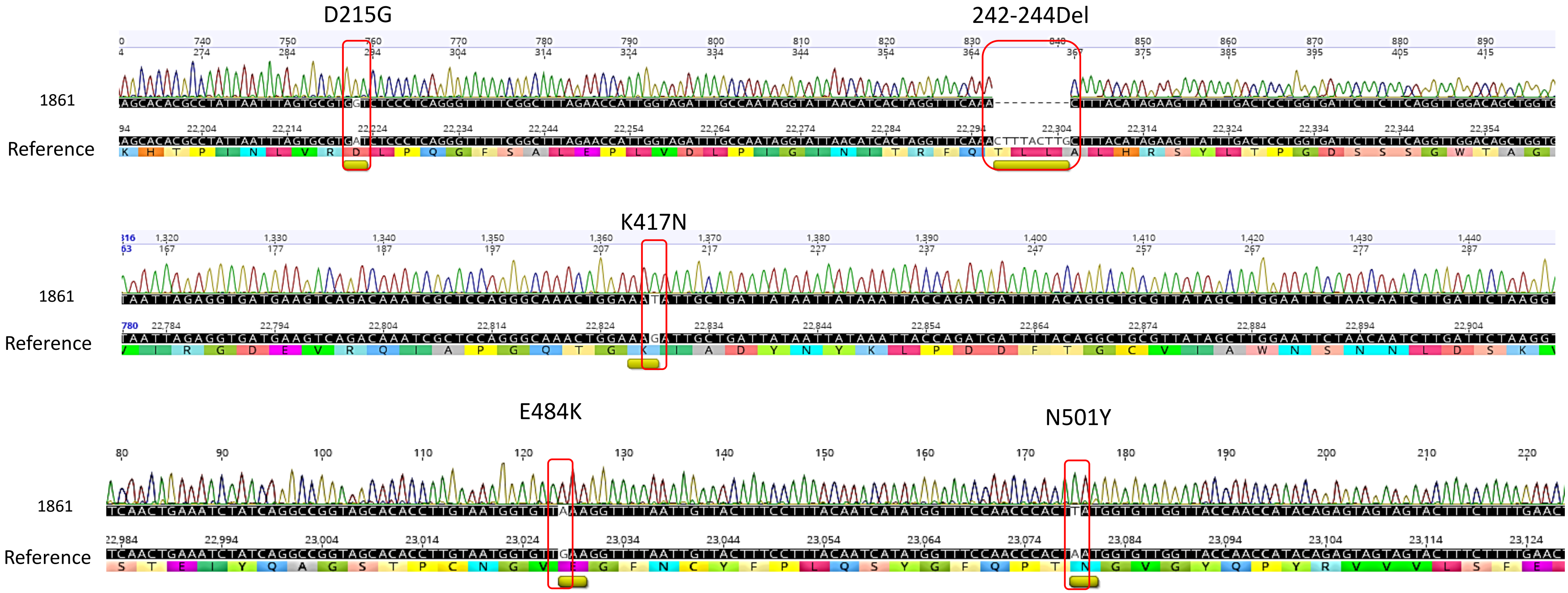


**<u>Supplementary Figure S6</u>.** Alignment of representative Spike gene N-terminal and RBD regions sequences of variant B.1.351 - suspected sample. The following mutations associated with variant B.1.351 were detected: D215G, 242-244del, K417N, E484K and N501Y. Mutations are marked with a rectangle and an underline under the sequence. Reference sequence: accession NC_045512.
